# The CRCbiome study: a large prospective cohort study examining the role of lifestyle and the gut microbiome in colorectal cancer screening participants

**DOI:** 10.1101/2020.12.22.20248658

**Authors:** Ane Sørlie Kværner, Einar Birkeland, Cecilie Bucher-Johannessen, Elina Vinberg, Jan Inge Nordby, Harri Kangas, Vahid Bemanian, Pekka Ellonen, Edoardo Botteri, Erik Natvig, Torbjørn Rognes, Eivind Hovig, Robert Lyle, Ole Herman Ambur, Willem M. de Vos, Scott Bultman, Anette Hjartåker, Rikard Landberg, Mingyang Song, Giske Ursin, Kristin Ranheim Randel, Thomas de Lange, Geir Hoff, Øyvind Holme, Paula Berstad, Trine B. Rounge

## Abstract

**Background:** Colorectal cancer (CRC) screening reduces CRC incidence and mortality. However, current screening methods are either hampered by invasiveness or suboptimal performance, limiting their effectiveness as primary screening methods. To aid in the development of a non-invasive screening test with improved sensitivity and specificity, we have initiated a prospective biomarker study (CRCbiome), nested within a large randomized CRC screening trial in Norway. We aim to develop a microbiome-based classification algorithm to identify advanced colorectal lesions in screening participants testing positive for an immunochemical fecal occult blood test (FIT). We will also examine interactions with host factors, diet, lifestyle and prescription drugs. The prospective nature of the study also enables the analysis of changes in the gut microbiome following the removal of precancerous lesions.

**Methods:** The CRCbiome study recruits participants enrolled in the Bowel Cancer Screening in Norway (BCSN) study, a randomized trial initiated in 2012 comparing once-only sigmoidoscopy to repeated biennial FIT, where women and men aged 50-74 years at study entry are invited to participate. Since 2017, participants randomized to FIT screening with a positive test result have been invited to join the CRCbiome study. Self-reported diet, lifestyle and demographic data are collected prior to colonoscopy after the positive FIT-test (baseline). Screening data, including colonoscopy findings are obtained from the BCSN database. Fecal samples for gut microbiome analyses are collected both before and 2 and 12 months after colonoscopy. Samples are analyzed using metagenome sequencing, with taxonomy profiles, and gene and pathway content as primary measures. CRCbiome data will also be linked to national registries to obtain information on prescription histories and cancer relevant outcomes occurring during the 10 year follow-up period.

**Discussion:** The CRCbiome study will increase our understanding of how the gut microbiome, in combination with lifestyle and environmental factors, influences the early stages of colorectal carcinogenesis. This knowledge will be crucial to develop microbiome- based screening tools for CRC. By evaluating biomarker performance in a screening setting, using samples from the target population, the generalizability of the findings to future screening cohorts is likely to be high.

**Trial Registration:** ClinicalTrials.gov Identifier: NCT01538550

## Background

Colorectal cancer (CRC) is a major global health burden, accounting for nearly 10% of all cancers diagnosed and cancer-related deaths each year (1). Although a decline in the age- standardized mortality rate has been observed over the past two to three decades in many countries (2–4), death rates remain high, particularly when diagnosed at later stages (5-year survival rate of 13% for metastatic disease compared to 90% when diagnosed at a localized stage) (1,5). The significant contribution to global cancer deaths, together with the worrying rise in incidence rates seen globally (3), especially the recent increase among younger age groups (6,7), highlights the need for widespread prevention strategies that are both effective and feasible on a large-scale basis.

There are two major precursor lesions of CRC: adenomatous polyps, accounting for the majority of cases, and serrated lesions, estimated to underlie up to 30% of CRC (8). The progression of precursor lesions to CRC is a long-term process, spanning a period of 10-15 years for most lesions (9). During this long latency period, most cancers develop asymptomatically, making them difficult to detect at a preclinical stage. Therefore, international guidelines recommend screening, with the aim of detection and removal of precancerous lesions to prevent cancer from occurring, or to detect cancer at the earliest stage possible (10–13).

Screening has been shown to reduce both CRC incidence (14–17) and mortality (14–21) in randomized controlled trials, even though current screening methods have known limitations (22). At present, the most commonly used screening method is the fecal immunochemical test (FIT) for occult blood, having mostly replaced the less sensitive guaiac-based fecal occult blood test (gFOBT) (23). Despite being more sensitive, performance characteristics are still suboptimal with regards to sensitivity and specificity, resulting in both missed neoplasms and unnecessary colonoscopy referrals (22). Of particular concern has been the limited performance in detecting precancerous lesions, representing a missed opportunity given the great potential for cancer prevention following removal of these lesions. There is also evidence that current screening methods perform worse for right-sided tumors, compared to left-sided ones (24), as well as in women compared to men (25,26). Thus, there is a requirement for screening methods and tools with improved performance for the entire screening population.

Both observational and experimental evidence point to an important role of the gut microbiome in development and progression of CRC (27). Numerous studies have demonstrated differences in the gut microbiome of tumor and adjacent non-tumor tissue (28,29), as well as in stool samples from CRC patients and healthy controls (30–38). Typically, the presence of a colorectal tumor has been associated with enrichment of pathogenic bacterial species, such as *Fusobacterium nucleatum, Escherichia coli* and *Bacteroides fragilis*, and depletion of potentially protective bacteria (e.g. producers of short chain fatty acid (SCFAs)) (27). Although less studied, there are reports indicating that subjects with precancerous lesions display shifts in their microbial profiles (30,33,39), suggesting the presence of microbial changes at early stages of colorectal carcinogenesis.

The gut microbiome is heavily influenced by the environment (40). Established risk factors for CRC, such as excess body weight, physical inactivity and a Western dietary pattern (typically high in red and processed meat and low in whole grains and dietary fiber) and protective factors, such as dairy products and use of certain medications (e.g. aspirin/NSAIDs and metformin) are suggested to modify the gut microbiome (41). At the same time, accumulating evidence indicates that modifications of the gut microbiome may allow environmental risk factors to induce malignant transformation (42,43). This highlights the complex relationship between the environment and the microbiome in the etiology of CRC.

The connection between a potentially pathogenic gut microbiome and CRC has resulted in a growing interest in the use of gut microbial biomarkers as screening tests for early detection of precancerous and cancerous lesions. Several studies have shown that combining microbiome data with the results of established screening methods, such as gFOBT or FIT, substantially increase the ability to classify groups of individuals with healthy colons, adenoma and CRC (30,33,34). Two recent meta-analyses of metagenome data showed that both taxonomic and functional gut microbial profiles predicted CRC at time of diagnosis with high accuracy (44,45).

Although results from previous biomarker studies are promising, no microbial biomarkers are currently used in national screening programs. In order to advance the utility of the gut microbiome in screening, additional data from prospective studies are needed.

## Objectives

The primary aim of the CRCbiome study is to develop a classification algorithm for identification of advanced colorectal lesions based on the screened individuals’ gut metagenome, demographics and lifestyle. Secondary aims are to provide a deeper understanding of how the gut microbiome evolves prior to a cancer diagnosis, as well as its interactions with host, lifestyle and environmental factors:

I. Identification of associations of the gut microbiome with advanced colorectal lesions, defined as presence of advanced adenomas, advanced serrated lesions or CRC, at baseline
II. Examination of interactions of the gut microbiome with host factors, diet, lifestyle and medication use on risk of advanced colorectal lesions at baseline
III. Description of changes in the gut microbiome following removal of precursor lesions of CRC

Long-term outcomes (i.e. incidence and mortality of advanced colorectal lesions) will be examined by means of passive follow-up using data from the national registries. The outcome assessment will be aligned with the 10 year follow-up of the Bowel Cancer Screening in Norway (BCSN) trial (46), from which the CRCbiome study recruits participants.

## Methods

### Study design

The CRCbiome study is a prospective cohort study nested within the BCSN trial, which is a pilot for a national screening program, organized by the Cancer Registry of Norway. The BCSN study is designed as a randomized trial comparing once-only sigmoidoscopy with FIT tests every two years for a maximum of four rounds (46). The trial was started in 2012, with follow-up FIT rounds scheduled to be completed in 2024. Participants randomized to the FIT group who test positive (i.e. hemoglobin >15 mcg/g feces), are referred for follow-up colonoscopy at their local screening center. Neoplastic lesions detected as part of the screening examination are removed during colonoscopy or elective surgery, if necessary. Biennial FIT testing is discontinued for those having undergone colonoscopy following a positive FIT test.

The CRCbiome study recruits participants from the BCSN trial who receive a positive FIT test. FIT positive participants are selected since they are referred to follow-up colonoscopies in line with the BCSN study protocol and will have detailed clinicopathological information. Conversely, as no diagnostic information is available for those with a negative FIT test, these are not included in the CRCbiome study. Of note, as recruitment for the CRCbiome study started five years after commencement of the BCSN trial, those with positive FIT findings in the first and initial part of the second round of screening in the BCSN were not invited. Even so, due to incomplete participation in the first round of FIT testing, 10% of the CRCbiome participants had their inclusion sample as their first screening test.

Participants are invited to the CRCbiome study prior to their colonoscopy examination. The invitation includes an information letter and two questionnaires (further details given below). FIT-positive fecal samples from the BCSN are retrieved following enrolment and represent the baseline sample of the CRCbiome study. Participants are thereafter contacted 2 and 12 months after colonoscopy for collection of follow-up fecal samples using the same sampling method. Fecal samples are processed for microbiome analysis as they become available to the project.

Based on the colonoscopy examination, participants are categorized into diagnostic groups ranging from no pathological findings to presence of advanced lesions and CRC. The groups selected for analyses will vary depending on aim (see <u>Outcome variables</u> for a complete description of outcomes).

Data collected in the CRCbiome study will be linked to national registries, including the Norwegian Prescription Database (47) and the Cancer Registry of Norway (48). An overview of the study design is shown in **Figure 1**. The design and handling of data in the CRCbiome study is in accordance with the STROBE guidelines for observational and metagenomics studies (49–51).

**Figure 1.**
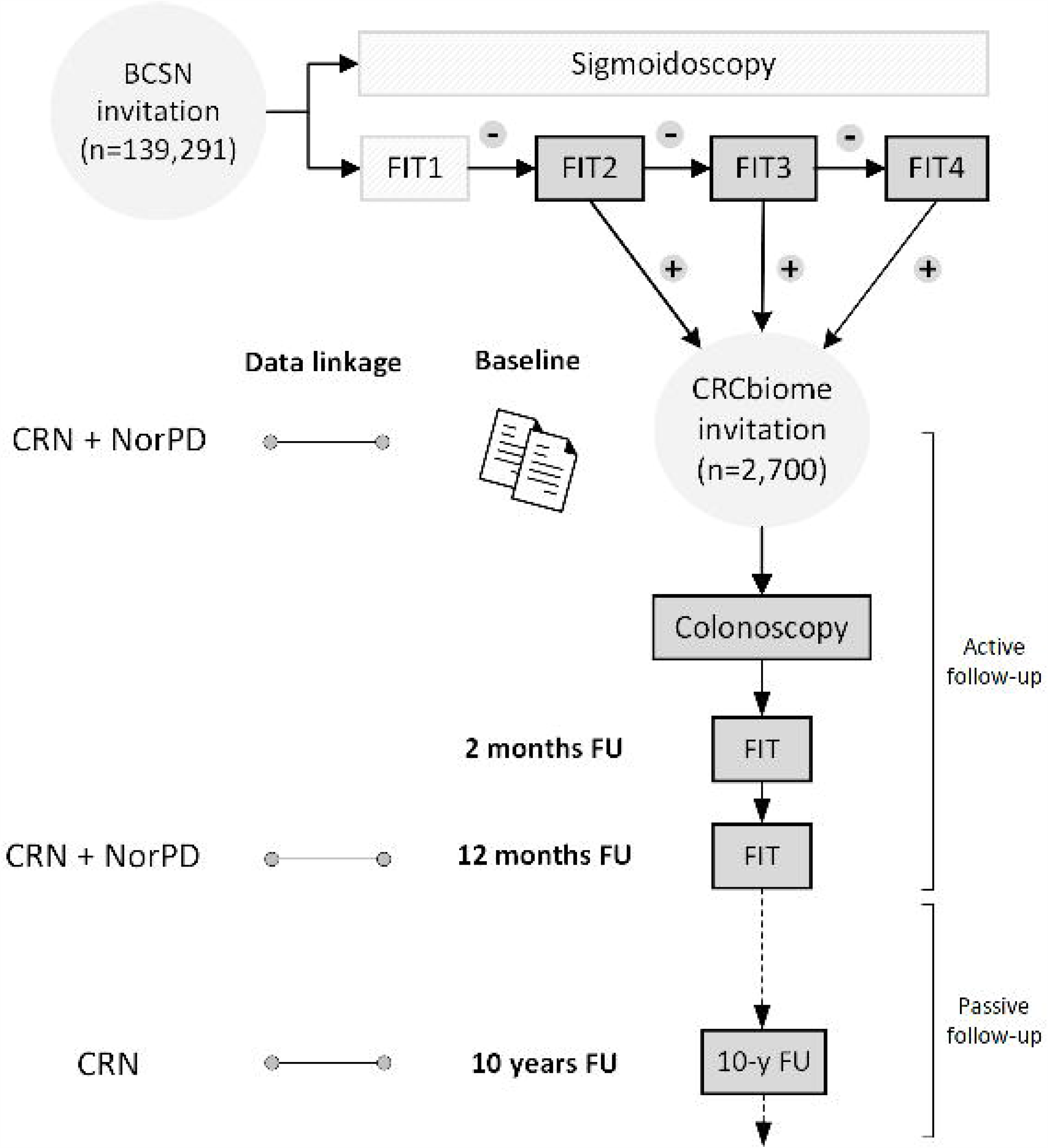
Flowchart of the CRCbiome study, nested within the BCSN. Abbreviations: BCSN, Bowel Cancer Screening in Norway; CRN, Cancer Registry of Norway; FIT, fecal immunochemical test; FU, follow-up; NorPD, Norwegian Prescription Database.

### Participants and eligibility

The BCSN trial includes 139,291 women and men aged 50-74 years in 2012, living in South- East Norway. Of these, 70,096 have been randomized to FIT screening. So far, the cumulative participation rate for the first three FIT rounds has been 68% (46). All screening participants with a positive FIT test are eligible for the CRCbiome study. Recruitment for the CRCbiome study started in 2017, and will continue until a minimum of 2,700 participants have been invited. So far, 2,426 have been invited and 1,413 (58%) have agreed to participate. With the current participation rate, we expect recruitment to be completed by March 2021 with a final number of participants of about 1,600 (see below for the sample size considerations).

The main inclusion and exclusion criteria for the BCSN trial and the CRCbiome study are listed in **Table 1**.

**Table 1.**
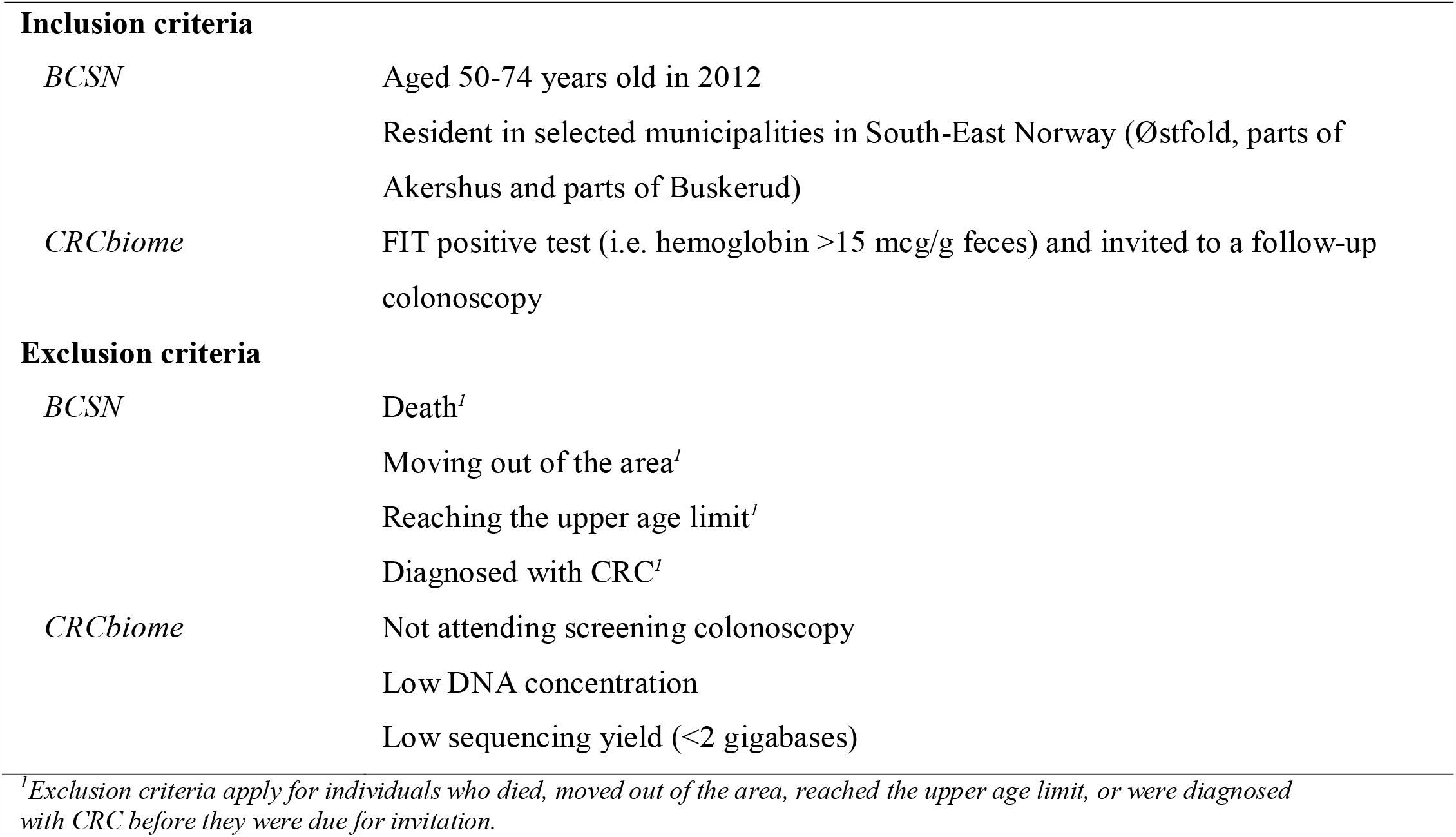
Inclusion and exclusion criteria in the BCSN trial and CRCbiome study.

Inclusion criteria
|  |  |
| --- | --- |
| *BCSN* | Aged 50-74 years old in 2012<br>Resident in selected municipalities in South-East Norway (Østfold, parts of Akershus and parts of Buskerud) |
| *CRCbiome* | FIT positive test (i.e. hemoglobin >15 mcg/g feces) and invited to a follow-up colonoscopy |
| Exclusion criteria |  |
| *BCSN* | Death<sup>1</sup><br>Moving out of the area<sup>1</sup><br>Reaching the upper age limit<sup>1</sup><br>Diagnosed with CRC<sup>1</sup> |
| *CRCbiome* | Not attending screening colonoscopy<br>Low DNA concentration<br>Low sequencing yield (<2 gigabases) |

<sup>1</sup>Exclusion criteria apply for individuals who died, moved out of the area, reached the upper age limit, or were diagnosed with CRC before they were due for invitation.

### Recruitment of participants

Eligible subjects are invited after being informed about their positive FIT test and a colonoscopy appointment has been scheduled. Invitations to the CRCbiome study, including the two questionnaires, are sent out by mail a minimum of four days prior to the colonoscopy. Returning at least one of the two questionnaires is regarded as a consent to the study, and includes permission to collect, analyze and store fecal samples, and to retrieve information from questionnaires and health registries.

Both the BCSN trial and the CRCbiome study have been approved by the Regional Committee for Medical Research Ethics in South East Norway (Approval no.: 2011/1272 and 63148, respectively). The BCSN is also registered at clinicaltrials.gov (Clinical Trial (NCT) no.: 01538550).

### Outcome variables

For the first two aims, the outcome variable will be defined based on the colonoscopy result. Participants will be grouped into four main categories: no confirmed neoplastic findings (Group 1); non-advanced lesions (Group 2); advanced lesions (Group 3); and CRC (Group 4) (**Table 2**). We may further subdivide lesions by clinicopathological features, including histopathological subtype (e.g. adenomas versus serrated lesions) and site of occurrence (proximal versus distal colon). Also of interest is the potential for distinct roles of environmental factors and the gut microbiome in the two main pathways of colorectal carcinogenesis: the adenoma-carcinoma pathway, and the serrated carcinoma pathway.

**Table 2.** Main outcomes of the screening colonoscopy among CRCbiome participants with preliminary distribution in percentages as of November 2020.

| **Colonoscopy result** | **Percentages<sup>1</sup>** |
| --- | --- |
| FIT+, no colonoscopy | 3.6 |
| **Group 1** |  |
| Negative | 11.2 |
| Polyp without histology<sup>2</sup> | 2.4 |
| Non neoplastic findings | 18.2 |
| **Group 2** |  |
| Non-advanced serrated lesions<sup>3</sup> | 6.4 |
| Non-advanced adenomas (<3) | 23.6 |
| Non-advanced adenomas (≥3) | 8.4 |
| **Group 3** |  |
| Advanced serrated lesions<sup>4</sup> | 4.4 |
| Advanced adenoma<sup>5</sup> | 18.1 |
| **Group 4** |  |
| CRC<sup>6</sup> | 3.6 |

<sup>1</sup>An extended version of this table, with colonoscopy result by FIT round, is shown in Additional file 1 (Supplementary table 2). In cases of multiple findings, participants are allocated to the most severe group. Numbers will therefore add up to 100%.
<sup>2</sup>Polyps lost during colonoscopy or where the endoscopist considers biopsy unnecessary, for example hyperplastic polyps in the rectum.
<sup>3</sup>Includes hyperplastic polyps with size <10 mm and sessile serrated lesions without dysplasia and size <10 mm.
<sup>4</sup>Defined as any serrated lesions with size ≥10 mm or dysplasia.
<sup>5</sup>Defined as any adenoma with either villous histology (≥25% villous components), high-grade dysplasia or polyp size greater than or equal to 10 mm (89).
<sup>6</sup>Defined as presence of adenocarcinoma arising from the colon or rectum. Collectively, advanced adenoma or CRC are referred to as advanced neoplasia (89).

For the third aim, the outcome variable will be defined based on the metagenome data. We will monitor several aspects of the gut microbiome, leveraging information derived from metagenomic sequencing to describe the presence of bacterial strains and the functional potential in paired samples during re-establishment of the gut microbiome following bowel cleansing and colonoscopy.

Long-term effects in the study will be assessed 10 years after recruitment is completed. This will include an investigation of incidence and mortality of advanced colorectal lesions.

### Clinical data, biological sampling and questionnaires

#### Assessment of clinical data

As part of the BCSN (46), participants are contacted by a study nurse prior to follow-up colonoscopy, to obtain information on medical history. This includes prior colonoscopies and CT colonographies, comorbidities, drug use, gastrointestinal symptoms, smoking habits, and body weight and height (**Table 3**). A variety of data are collected in relation to the follow-up colonoscopy, including screening outcomes (i.e. presence and clinicopathological characterization of detected lesions) and cgaracteristics relevant to the endoscopic procedure (**Table 3**). For all lesions detected, size, location, appearance, technique used for removal and tissue sampling, and completeness of removal are recorded. Both the medical history data and data collected as part of the follow-up colonoscopy are entered into a dedicated database by the responsible health care provider. A complete overview of the data collected in the BCSN trial can be found elsewhere (46).

**Table 3.** Data sources and output generated in the CRCbiome study.

|  |  | Time points<sup>1</sup> |  |  |  |
| --- | --- | --- | --- | --- | --- |
|  |  | Baseline | 2 months | 12 months | 10 years |
| **Clinical data** |  |  |  |  |  |
| Medical history | Prior colonoscopies and CT colonographies, comorbidities (3 items), drug use (6 items), gastrointestinal symptoms (9 items), smoking habits (1 item) and body weight and height | x |  |  |  |
| Screening specific data | FIT value, endoscopic findings, histopathology and clinical diagnoses, type of procedure and bowel preparation used, degree of bowel cleansing, intubation level, duration of colonoscopy, use of sedation or analgesia, reason for ending the examination, if necessary, and recommended surveillance | x |  |  |  |
| **Biological samples** |  |  |  |  |  |
| Fecal samples | Gut microbiome profile, including taxonomic and functional profiles | x | x | x |  |
| **Questionnaires** |  |  |  |  |  |
| Lifestyle and demographics (LDQ) | Demographic factors (i.e. national background, marital status, education and occupation), smoking and snus habits (up to 5 questions each), physical activity (hours spent on physical activity of light, moderate and high intensity per week and presence of chronic diseases restricting ability of being physically active), use of regular and cultured milk (two frequency questions), mode of delivery at birth, removal of the appendix, recent use of medications (i.e. antibiotic and antacid usage the last three months), presence of chronic bowel disorders and food intolerances (closed and open format questions) and presence of CRC among first-degree relatives | x |  |  |  |
| Diet (FFQ) | Energy intake, intake of macro and micronutrients, frequency and/or amounts of 256 foods and drinks consumed<sup>2</sup>, dietary patterns, including meal pattern, body weight and height | x |  |  |  |
| **Registry data** |  |  |  |  |  |
| Cancer registry | Cancer incidence and mortality, clinicopathological characteristics, information on treatment regimens | x |  | x | x |
| Prescriptions database | Complete prescription history since 2004 | x |  | x |  |

<sup>1</sup>In cases of multiple screening colonoscopies, the time of the 2 and 12 months follow-up visits is defined based on the first and last colonoscopy, respectively.
<sup>2</sup>A complete overview of the food items included in the FFQ is given in Additional file 1 (Supplementary table 1).

#### Biological sampling and gut microbiome analysis

##### FIT sampling and storage

Sampling kits for stool sample collection are mailed to the participants three times during the study period. No restrictions on diet or medication use are required prior to sampling. Stool is collected using plastic sticks, which collect about 10 mg stool. The stool is then stored in 2 ml of buffer containing HEPES (4-(2-hydroxyethyl)-1-piperazineethanesulfonic acid), BSA (Bovine serum albumin) and sodium azide. Samples are then packed in padded envelopes and returned by mail to a laboratory at Oslo University Hospital for analysis and further storage at -80 °C. Shipping time is estimated to 3–10 days. Immunochemical testing for blood in feces is performed continuously using the OC-Sensor Diana (Eiken Chemical, Tokyo, Japan) as samples are received at the laboratory.

##### DNA extraction

Thawed samples are transferred to three 500 ml aliquots from the sampling bottle using a blood sampling needle (Vacuette) perforating the plastic lid. Samples are stored at -80 °C until further processing.

Extraction of DNA is carried out using the QIAsymphony automated extraction system, using the QIAsymphony DSP Virus/Pathogen Midikit (Qiagen), after an off-board lysis protocol with some modifications. Each sample is lysed with bead-beating: a 500 µl sample aliquot is transferred to a Lysing Matrix E tube (MP Biomedicals) and mixed with 700 µl phosphate-buffered saline (PBS) buffer. The mixture is then shaken at 6.5 m/s for 45 s. After the bead-beating, 800 µl of the sample is mixed with 1055 µl of off-board lysis buffer (proteinase K, ATL buffer, ACL buffer and nuclease-free water) as recommended by Qiagen. The sample is incubated at 68 °C for 15 min for lysis. Nucleic acid purification is performed on the QIAsymphony extraction robot using the Complex800_OBL_CR22796_ID 3489 protocol, a modified version of the Complex800_OBL_V4_DSP protocol. Purified DNA is eluted in 60 µl AVE-buffer (Qiagen). DNA purity is assessed using a Nanodrop2000 (Thermo Fisher Scientific, USA), and the concentration is measured by Qubit (Thermo Fisher Scientific, USA).

##### Metagenome sequencing

Libraries for metagenome sequencing are prepared from extracted DNA at the sequencing laboratory of the Institute for Molecular Medicine Finland FIMM Technology Centre, University of Helsinki (P.O. Box 20, University of Helsinki, Finland) using Illumina sequencing, with the aim of producing 3 gigabases of DNA sequence per sample.

In details, 29 µl of extracted DNA is purified and concentrated by adding an equal volume of AMPure XP (Beckman Coulter Life Sciences, Indianapolis, IN, USA). Purification is then performed as per the manufacturer’s instructions. The purified samples are eluted to 17 µl of 10 mM Tris-HCl, pH 8.5, and DNA concentrations are determined by Quant-iT PicoGreen dsDNA Assay Kit (Thermo Fisher Scientific, Waltham, MA, USA). The samples are normalized to a maximum concentration of 3.3 ng/µl, resulting in DNA inputs of 25 ng or less.

Sequencing libraries are prepared according to the Nextera DNA Flex Library Prep Reference Guide (v07) (Illumina, San Diego, CA, USA), with the exception that the reaction volumes are scaled down to ¼ of the protocol volumes. The libraries are amplified according to the protocol with 7 PCR cycles. All the library preparation steps are performed on a Microlab STARlet (Hamilton Company, Reno, NV, USA) and Biomek NX□ (Beckman Coulter Life Sciences, Indianapolis, IN, USA) liquid handlers running custom scripts.

DNA concentrations of the finished libraries are determined with Quant-iT PicoGreen dsDNA Assay. Libraries are combined into pools containing 240 libraries with 4.5 ng of each library using Echo 525 Acoustic Liquid Handler (Beckman Coulter Life Sciences, Indianapolis, IN, USA). Library pools are size-selected to a fragment size range between 650 and 900 bp using BluePippin (Sage Science Beverly, MA, USA).

Sequencing is performed with the Illumina NovaSeq system using S4 flow cells with lane divider (Illumina, San Diego, CA, USA). Each pool is sequenced on a single lane. Read length for the paired-end run is 2×151 bp.

##### Processing and analysis of sequencing data

Sequencing data are transferred to a platform for secure storage and analysis of sensitive research-related data at the University of Oslo (52). The analysis of metagenomic sequencing data is handled in a uniform manner using a customizable workflow manager (53). To establish a quality-filtered dataset, standard filters are applied: sequences corresponding to adapters used in library preparation, being of low quality (54) and those mapping to the human genome (55), with subsequent quality control of filtered sequencing reads (56).

Taxonomic classification and determination of microbial gene content, including functional annotation (e.g. using gene ontology and KEGG databases) will be performed using publicly available tools. Abundance measures will be used to calculate taxonomic and functional alpha and beta diversity, as well as serving as input for machine learning approaches aimed at producing classifiers for high-risk individuals in a data-driven manner. Further metagenome-derived measures may include identification of metagenome-assembled genomes, strain-level analysis and description of the gut virome.

#### Questionnaires

Two questionnaires are used to collect data on diet, lifestyle and demographic data: a food frequency questionnaire (FFQ) and a general lifestyle and demographics questionnaire (LDQ). Self-reported dates of questionnaire completion are registered in the project database. Returned questionnaires are reviewed manually before scanning and further processing. In cases of low-quality data, participants are contacted for clarification.

##### Assessment of dietary intake

Dietary intake is assessed using a semiquantitative, 14-page FFQ, designed to assess the habitual diet during the preceding year. The questionnaire is a modified version of an FFQ developed and validated by the Department of Nutrition, University of Oslo (57–62). The questionnaire has been validated for both energy intake (57–59), intake of macro and micronutrients (57,59,62), as well as selected food items and groups (59–62). The questionnaire includes 23 main questions, covering a total of 256 food items, as well as a free-text field for entries of food items not covered by the questionnaire. For each food item (except one on preferred types of fat for cooking), participants are asked to record frequency of consumption, ranging from never/seldom to several times a day, and/or amount, typically as portion size given in various household units (e.g. deciliters, glasses, cups, spoons). In total, there are 249 questions on frequency, 204 on portion size, one on preferences and nine other, mostly related to meal patterns (**Additional file 1, supplementary Table 1**).

As with any dietary assessment method, the FFQ is prone to errors due to inaccurate reporting and missing answers. Therefore, to mitigate such errors, a standardized framework for how to review and evaluate FFQ quality has been developed. A detailed overview of the framework is given in **Additinoal file 2, supplementary Figure 1**. In brief, incoming FFQs are reviewed by trained personnel according to a set of predefined criteria. Scanning of questionnaires is performed using the Cardiff TeleForm program (Datascan, Oslo, Norway). The dietary calculation system KBS (short for “**K**ost**b**eregnings**s**ystem”), developed at the Department of Nutrition, University of Oslo, is used to calculate food and nutrient intake. The latest version of the food database (i.e. AE-18 or newer) will be used, which is largely based on the Norwegian Food Composition Table (63). In line with common practice in nutrition studies, missing answers are imputed as zero intake (59,61,64,65) and observations with extreme energy intake levels in both the upper and lower range will be excluded (66).

The main focus of the dietary analyses will be on foods and drinks linked to the risk of CRC and its precursor lesions, including intakes of alcohol, red and processed meat, wholegrains, foods containing dietary fiber, dairy products and calcium supplements (67). Dietary intake will also be studied holistically by employing various dietary indices such as the 2018 World Cancer Research Fund/American Institute for Cancer Research (WCRF/AICR) index for adherence to cancer prevention recommendations (68).

##### Assessment of lifestyle and demographic data

Lifestyle and demographic data are assessed using a four page questionnaire, based on questions used in previous national surveys (69,70). Prior to the study start, the questionnaire was piloted in a targeted population and adjusted based on feedback from pilot study participants. The questionnaire has ten main questions, covering demographic factors (education, occupation and marital status), diagnosis of CRC among first-degree relatives, presence of chronic bowel disorders and food intolerances, removal of the appendix, mode of delivery at birth, smoking and snus (i.e. smokeless tobacco) habits, recent use of medications, the past years’ physical activity level and lastly use of regular and cultured milk, which is not completely covered in the FFQ (see **Table 3** for a detailed overview). In the questions concerning smoking and snus habits, participants are asked to recall their current habits, including the daily number of cigarettes/snus portions, as well as years since possible cessation and total years of use. Questionnaires are scanned and processed using the Cardiff TeleForm program (InfoShare, Oslo, Norway).

### Registry data

Data collected in the CRCbiome study will be linked to national registries, including the Norwegian Prescription Database and the Cancer Registry of Norway, using personal identification numbers. Complete data linkages will be undertaken twice during active follow-up: after all participants have completed baseline and diagnostic information from follow-up colonoscopies is available, and then after the one-year follow-up is completed. In addition, linkage to the Cancer Registry of Norway will be performed at least once during the 10 year follow-up period.

#### Norwegian Prescription Database

The Norwegian Prescription Database (71) will be used to obtain information on medication history prior to CRC screening, and during the first year of follow-up. The registry contains data on all medications prescribed to Norwegian citizens since 2004. Prescription drugs are categorized according to the Anatomical Therapeutic Chemical (ATC) system, a hierarchical classification system developed by the WHO (72,73). For each drug, the number of packages dispensed, the number of defined daily doses (DDD), the prescription category, and the date of dispensing are registered.

Linkage to the Norwegian Prescription Database enables an in-depth analysis of associations between drug use, the gut microbiome and advanced colorectal lesions. Initially, we will perform drug-wide association analyses to screen for potential associations, adjusting for key covariates. Detected associations will then be examined in detail, including a more refined categorization of drug variables, robust covariate adjustments as well as the analysis of timing and dose-response relations. Prescription histories will also be used as a proxy for life-long burden of chronic diseases.

#### Cancer Registry of Norway

Information on clinicopathological characteristics, cancer therapy, as well as outcomes assessed as part of passive follow-up, will be obtained from the Cancer Registry of Norway (74). The Cancer Registry of Norway has recorded incident cancer cases on a nationwide basis since 1953 and has been shown to have accurate and almost complete ascertainment of cases (98.8% for the registration period 2001-2005) (75). According to recent estimates, about 93% of all cancer cases and ≥95% of cancers in the colon and rectum are morphologically verified (48). Cancer diagnoses are recorded using the International Classification of Diseases, version 10 (ICD-10). Mortality data in the registry are obtained from the Cause of Death Registry and coded using the same ICD-10 categories as for the incidence data.

### Data processing and management

To facilitate project administration, including recruitment and follow-up of participants, custom software has been developed. This application communicates with two project specific databases (i.e. the BCSN and CRCbiome databases). Only authorized data manager personnel have complete access to the datasets. A simplified version of the data generation process is depicted in **Figure 2**.

**Figure 2.**
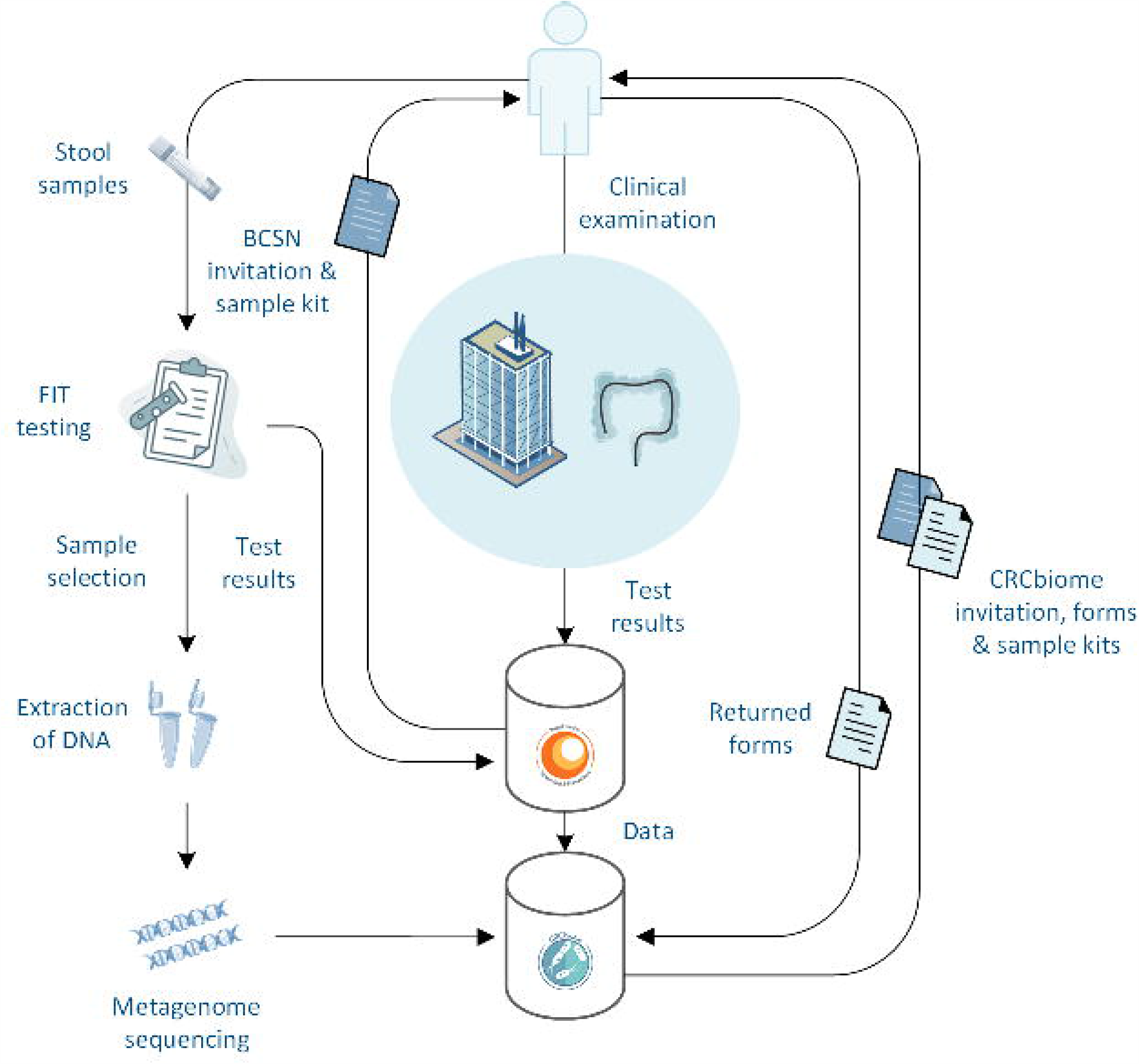
Simplified version of the data generation process in CRCbiome. The figure is created based on free images from Servier Medical Art (Creative Commons Attribution Liscence, creativecommons.org/liscences/by/3.0/) and Stockio (https://www.stockio.com/).

In line with common practice for linkage with national registries (76), linked data will receive unique ID numbers specific to the particular project. Linkage of research data will be performed by the data controller. For the metagenome data, which due to its size cannot be transferred using ordinary methods, linkage will be performed in-house by an independent data manager without access to other parts of the data than those strictly necessary for linkage.

All data collected in the CRCbiome study will be stored and analyzed at a platform for secure handling of sensitive research-related data, operated by the University of Oslo (52). Access to research data for external investigators, or use outside of the current protocol, will require approval from the Norwegian Regional Committee for Medical and Health Research Ethics and a data access committee (information available on the project web site (77)). Research data are not openly available because of the principles and conditions set out in articles 6 (1) (e) and 9 (2) (j) of the General Data Protection Regulation (GDPR).

### Sample size considerations

The number of participants to include was chosen with the aim of providing adequate power for the development of a highly sensitive classification algorithm via data-driven analyses of gut metagenomes that will accurately identify FIT-positive individuals in need of clinical intervention.

The classifier will be trained using counts of taxonomic units, signature and genes categorized according to gene ontology or pathway membership from metagenomes, FFQ, demographic and lifestyle data as input variables, and advanced colorectal lesions as outcome (i.e. group group 3 and group 4, **Table 2**). The CRC risk classification will be done using machine learning algorithms suited to metagenome data, such as lasso regression (78), support-vector machines (79), random forests (80), multi-layer perception neural networks (81) and scalable tree boosting (82) algorithms. Evaluation of the classifier will be conducted in a leave-out test set. As outlined below, we believe that with sufficient sample size, development of a classifier with a sensitivity of 0.95 is achievable in the training set, being within the range of published reports (30,33).

With a projected classifier sensitivity of 0.95 and a minimally acceptable sensitivity of 0.8, at 80% power and 95% confidence level, 50 participants with advanced colorectal lesions are required in the test set (83). Classifier specificity in the setting of FIT-positive individuals will have a lower requirement, and we therefore set the expected classifier specificity to 0.75 and a minimally acceptable specificity of 0.6, thus requiring 100 participants with normal findings in the test set. Based on initial recruitment, we expect a participation rate of 58%, with 26% of participants having findings of advanced lesions or CRC (**Table 2**). By inviting 2,700 FIT-positive BCSN participants, and splitting the training and test sets 80/20, a projected number of 1,253 and 313 participants will constitute the training and test sets, respectively, which will include adequate numbers of participants with both advanced colorectal lesions and normal findings in the test set. With this sample size, we will also be able to perform stratified analyses.

## Discussion

CRC remains a major public health challenge with substantial personal and societal costs (22). Screening is an effective measure to reduce disease burden (22). However, current screening methods suffer from limitations, limiting the number of preventable cases. Innovative use of currently available methods represents a promising avenue for improvements in CRC prevention (22). The current study is designed to contribute to the development of microbial biomarkers, using metagenome sequencing and comprehensive questionnaire and registry data for improved detection of advanced lesions and CRC in a FIT-positive population. The CRCbiome study is unique in that it uses data from the screening population to develop relevant biomarkers.

The idea of using microbial biomarkers to increase the performance of CRC screening has received increased attention with the adoption of high-throughput characterization of the gut microbiome. Ideally, combining microbial biomarkers with FIT testing could achieve the sensitivity of direct visualization methods and the uptake of non-invasive fecal tests. Several studies have demonstrated improved ability to discriminate individuals with healthy colons from those with advanced neoplasia when adding microbial biomarkers in the prediction model, more so for carcinoma (area under the curve (AUC) of 0.87-0.97 (30,33,34)) than adenoma (AUC of 0.76 (33)). Despite great promise, these studies have typically been limited by small sample sizes (30,32–34), cross-sectional designs (30–34), use of suboptimal or low-resolution methods to study the gut-microbiome (30–33) and lack of data on important confounders (30–34). The CRCbiome study seeks to address several of these shortcomings.

Major strengths of the CRCbiome study include its large sample size and prospective nature, use of state of the art methodology for studying the gut microbiome and access to detailed information on likely confounders of the relationship between the gut-microbiome and advanced colorectal lesions. A further strength of the study is in its organization and logistics structure, being nested within the BCSN. The immediate availability of clinically verified outcome data, via follow-up colonoscopies and cancer registry data, allow for prospective investigations on multiple outcomes relevant to the screening population (e.g. polyp recurrence). Access to comprehensive high-quality data on diet and lifestyle, including complete prescription histories, also enables the investigation of the predictive performance of more broad classifiers, laying the ground for personalized screening strategies, including risk-stratified approaches.

With a study population solely consisting of FIT positive participants, the projected number of individuals with high-risk lesions or CRC is relatively high (about 409 (26%), group 3 and 4, **Table 2**), thereby increasing the power to achieve accurate classification of advanced colorectal neoplasms. Still, whether findings in this population extends to cases missed by FIT testing is unknown.

Collection of follow-up samples at 2 and 12-months post colonoscopy represents an extension of the cross-sectional design of most prior studies, shedding light on the development of the gut microbiome following colonoscopy with or without removal of CRC precursor lesions. While there are examples of shifts in microbial profiles following colonoscopy, the gut microbiome typically reverts to the initial state within weeks (84). Deviations from re-establishment of the gut microbiome both in the medium and long term have the potential for causal interpretations.

The study also has some limitations. Exclusive selection of FIT positive participants may limit the generalizability of the findings to those with bleeding neoplastic lesions. Consequently, improvements in diagnostic performance may be limited to specificity, and thus the ability to correctly classify healthy individuals. However, since lesions tend to bleed intermittently (85) and the study aims to identify potential causal pathways, we consider it likely that the identified biomarker also may have improved sensitivity in the screening population as a whole.

A further limitation is the lack of information on fecal metrics such as the Bristol stool scale, which has been shown to be an important determinant of microbiota richness and variance (86). However, variation in microbiome profile due to stool consistency could likely be explored by use of gastrointestinal symptoms as a surrogate, data on which is available in the BCSN database.

Lastly, lack of follow-up data on diet and lifestyle may complicate the interpretation of microbial changes following colonoscopy. Even though prior studies in comparable study populations show that potential changes in diet and lifestyle following screening are modest (87,88), caution in interpretation of follow-up samples is warranted.

The CRCbiome study represents a valuable source of data for further research. An example is access to complete prescription histories from the Norwegian Prescription Database that enables in-depth analyses of associations between a broad range of medications, microbial features and neoplasia risk, both during short and long-term follow-up. The fecal samples collected are also biobanked and can be used for other purposes beside the study aims of the current protocol. For instance, in addition to metagenome sequencing, the fecal samples can potentially be used for other omics analyses, such as transcriptome and metabolome analysis. All tissue specimens removed during colonoscopy are also available to the project, enabling in-depth molecular profiling.

## Conclusion

The CRCbiome study investigates the role of the gut microbiome, and its interactions with host factors, diet and lifestyle, in early stage colorectal carcinogenesis. Information obtained from this project will guide the development of a microbial biomarker for accurate detection of advanced colorectal lesions. By performing biomarker discovery within a screening population, the generalizability of the findings to future screening cohorts is likely to be high.

## Data Availability

Due to the principles and conditions set out in articles 6 (1) (e) and 9 (2) (j) of the General Data Protection Regulation (GDPR), research data generated in the CRCbiome study are not openly available. Further information on access to CRCbiome data can be found on the project web site: https://www.kreftregisteret.no/en/Research/Projects/microbiota-and-lifestyle-in-colorectal-cancer-screeing/

## List of abbreviations

AICR: American Institute for Cancer Research
ATC: Anatomical Therapeutic Chemical
AUC: area under the curve
BCA: bovine serum albumin
BCSN: Bowel Cancer Screening in Norway
Bp: base pair
CRC: colorectal cancer
CRN: Cancer Registry of Norway
CT: computed tomography
DDD: defined daily doses
DNA: deoxyribonucleic acid
DPIA: Data Processing Impact Assessment
FFQ: food frequency questionnaire
FIMM: Institute for Molecular Medicine Finland
FIT: fecal immunochemical test
FU: follow-up
gFOBT: guaiac-based fecal occult blood test
ICD: International Classification of Diseases
KBS: Kostberegningssystem (“Dietary calculation system”)
KEGG: Kyoto Encyclopedia of Genes and Genomes
LDQ: lifestyle and demographic questionnaire
NCT: National Clinical Trial
NorPD: Norwegian Prescription Database
NSAIDs: non-steroid anti-inflammatory drugs
PCR: polymerase chain reaction
PBS: phosphate-buffered saline
SCFA: short chain fatty acid
STROBE: Strengthening the Reporting of Observational Studies in Epidemiology
WCRF: World Cancer Research Fund
WHO: World Health Organization

## Declarations

### Ethics approval and consent to participate

The CRCbiome study is approved by the Norwegian Regional Committees for Medical and Health Research Ethics (Approval no.: 63148). Written informed consent to participate has been obtained from all participants at study enrollment. All biological materials are stored in a biobank at Oslo University Hospital.

### Consent for publication

Not applicable.

### Availability of data and materials

Due to the principles and conditions set out in articles 6 (1) (e) and 9 (2) (j) of the General Data Protection Regulation (GDPR), research data generated in the CRCbiome study are not openly available. Further information on access to CRCbiome data can be found on the project web site (77)).

### Competing interests

There are no competing interests.

### Funding

This project would not have been possible without funding from the Norwegian Cancer Society, the Research Council of Norway and the South Eastern Norway Regional Health Authority.

### Authors’ contributions

ASK and EB (Birkeland) had the main responsibility for writing the manuscript.

PB and TBR are the principal investigators.

All authors contributed to the study design and protocol.

All authors contributed to the writing and approval of the final manuscript

## Acknowledgements

We would like to aknowledge the devoted secretaries, nurses and doctors at Bærum and Moss hospital, and the biomedical laboratory scientists at Oslo University Hospital for their contributions to this study. We would also like to thank the personnel involved in sequencing all CRCbiome samples at the Sequencing laboratory of Institute for Molecular Medicine Finland FIMM Technology Centre, University of Helsinki. Lastly, we would like to thank each study participoant, as well as all collaborative partners, technicians and students that have, and will, contribute to this study.

## Additional files

Additional file 1: Supplementary tables (Additional file 1)

*Additional_file-1*.*docx*

Contains two supplementary tables.

Additional file 2: Supplemetary figures (Additional file 2)

*Additional_file-2*.*docx*

Contains a supplementary figure with figure title and legend.

Additional file 3: Ethical approval

*Additional_file-3*.*pdf*

A translated version of the ethical approval for the CRCbiome study.

Additional file 4: Funding

*Additional_file-4A*.*pdf*

*Additional_file-4B*.*pdf*

*Additional_file-4C*.*pdf*

The respective files contains documentation of funding (translated from Norwegian) from the Norwegian Cancer Society (4A-B) and the South Eastern Norway Regional Health Authority (4C). Documentation of funding from the Research Council of Norway can be submitted afterwords if required. The funding was provided to a large collabrotative project where the CRCbiome study only represented one part/work package.

Additional file 5: STROBE chechlist

*Additional_file-5*.*pdf*

Contains the STROBE checklist for observational studies.

## Notes

### Competing Interest Statement

The authors have declared no competing interest.

### Clinical Trial

NCT01538550

### Author Declarations

Both the BCSN trial, which the study recruits participants from, and the CRCbiome study have been approved by the Regional Committee for Medical Research Ethics in South East Norway (Approval no.: 2011/1272 and 63148, respectively). The BCSN is also registered at clinicaltrials.gov (Clinical Trial (NCT) no.: 01538550).

