## Additional file 1 for "The CRCbiome study: a large prospective cohort study examining the role of lifestyle and the gut microbiome in colorectal cancer screening participants"

**Additional file 1: Supplementary tables**

| **Supplementary Table 1**. Questions included in the Food Frequency Questionnaire (FFQ). | | | | |
| --- | --- | --- | --- | --- |
|  | **Types of questions** | | |  |
| **Questions (number of food items)** | Frequency | Portion size | Preferences | Other |
| 1. Bread (5 items) | 5 |  |  |  |
| 2. Butter/margarine on bread (11items) | 11 | 1 |  |  |
| 3. Spreads (30 items) | 30 |  |  |  |
| 4. Cereals (8 items) | 8 | 8 |  |  |
| 5. Jam on cereals | 2 | 2 |  |  |
| 6. Milk (8 items) | 8 |  |  |  |
| 7. Yoghurt (5 items) | 5 | 5 |  |  |
| 8. Cold drinks (non-alcoholic) (12 items) | 12 | 12 |  |  |
| 9. Alcoholic beverages (8 items) | 8 | 8 |  |  |
| 10. Hot drinks (non-alcohol) (10 items) | 10 | 10 |  |  |
| 11. Sugar/artificial sweeteners in coffee/tea (6 items) |  | 6 |  |  |
| 12. Dishes (47 items) | 47 | 47 |  |  |
| 13. Potatoes, rice, spaghetti and vegetables (25 items) | 25 | 25 |  |  |
| 14. Sauces and dressings (17 items) | 17 | 17 |  |  |
| 15. Preferred types of fat for cooking (1 item) |  |  | 1 |  |
| 16. Fruits and berries (17 items) | 17 | 17 |  |  |
| 17. Daily portions of fruits and vegetables (0 items) |  | 2 |  |  |
| 18. Desserts, cakes and snacks (27 items) | 27 | 27 |  |  |
| 19. Dietary supplements (17 items) | 17 | 17 |  |  |
| 20. Consumption pattern (0 items) |  |  |  | 5 |
| 21. Sex (0 items) |  |  |  | 1 |
| 22. Age (0 items) |  |  |  | 1 |
| 23. Weight and height (0 items) |  |  |  | 2 |
| **Total** | **249** | **204** | **1** | **9** |

| **Supplementary Table 2**. Main outcomes of the screening colonoscopy for CRCbiome participants in percentages as of November 2020, stratified by FIT round^1^. | | | | |
| --- | --- | --- | --- | --- |
|  | **FIT round** | | |  |
| **Colonoscopy result** | **2nd round** | **3rd round** | **4th round** | **Overall** |
| FIT+, no colonoscopy | 2.9 | 3.7 | 4.5 | 3.6 |
| **Group 1** |  |  |  |  |
| Negative | 11.7 | 11.8 | 9.1 | 11.2 |
| Polyp without histology^2^ | 0.2 | 2.4 | 5.6 | 2.4 |
| Non neoplastic findings | 18.7 | 17.8 | 18.5 | 18.2 |
| **Group 2** |  |  |  |  |
| Non-advanced serrated lesions^3^ | 5.6 | 6.4 | 7.3 | 6.4 |
| Non-advanced adenomas (<3) | 22.4 | 24.1 | 24.1 | 23.6 |
| Non-advanced adenomas (≥3) | 8.8 | 8.4 | 8.0 | 8.4 |
| **Group 3** |  |  |  |  |
| Advanced serrated lesions^4^ | 5.6 | 4.1 | 3.5 | 4.4 |
| Advanced adenoma^5^ | 21.4 | 16.9 | 16.1 | 18.1 |
| **Group 4** |  |  |  |  |
| CRC^6^ | 2.7 | 4.4 | 3.1 | 3.6 |

*^1^In cases of multiple findings, participants are allocated to the most severe group. Numbers will therefore add up to 100%.*

*^2^Polyps lost during colonoscopy or where the endoscopist considers biopsy unnescessary, for example hyperplastic polyps in the rectum.*

^3^Includes hyperplastic polyps with size <10 mm and sessile serrated lesions without dysplasia and size <10 mm.

^4^Defined as any serrated lesions with size ≥ 10 mm or dysplasia.

^5^Defined as any adenoma with either villous histology, high-grade dysplasia or polyp size equal to or greater than 10 mm (1).

^6^Defined as presence of adenocarcinoma arising from the colon or rectum. Collectively, advanced adenoma or CRC are referred to as advanced neoplasia (1).
