## Additional file 2 for "The CRCbiome study: a large prospective cohort study examining the role of lifestyle and the gut microbiome in colorectal cancer screening participants"

**Additional file 2: Supplementary Figures**

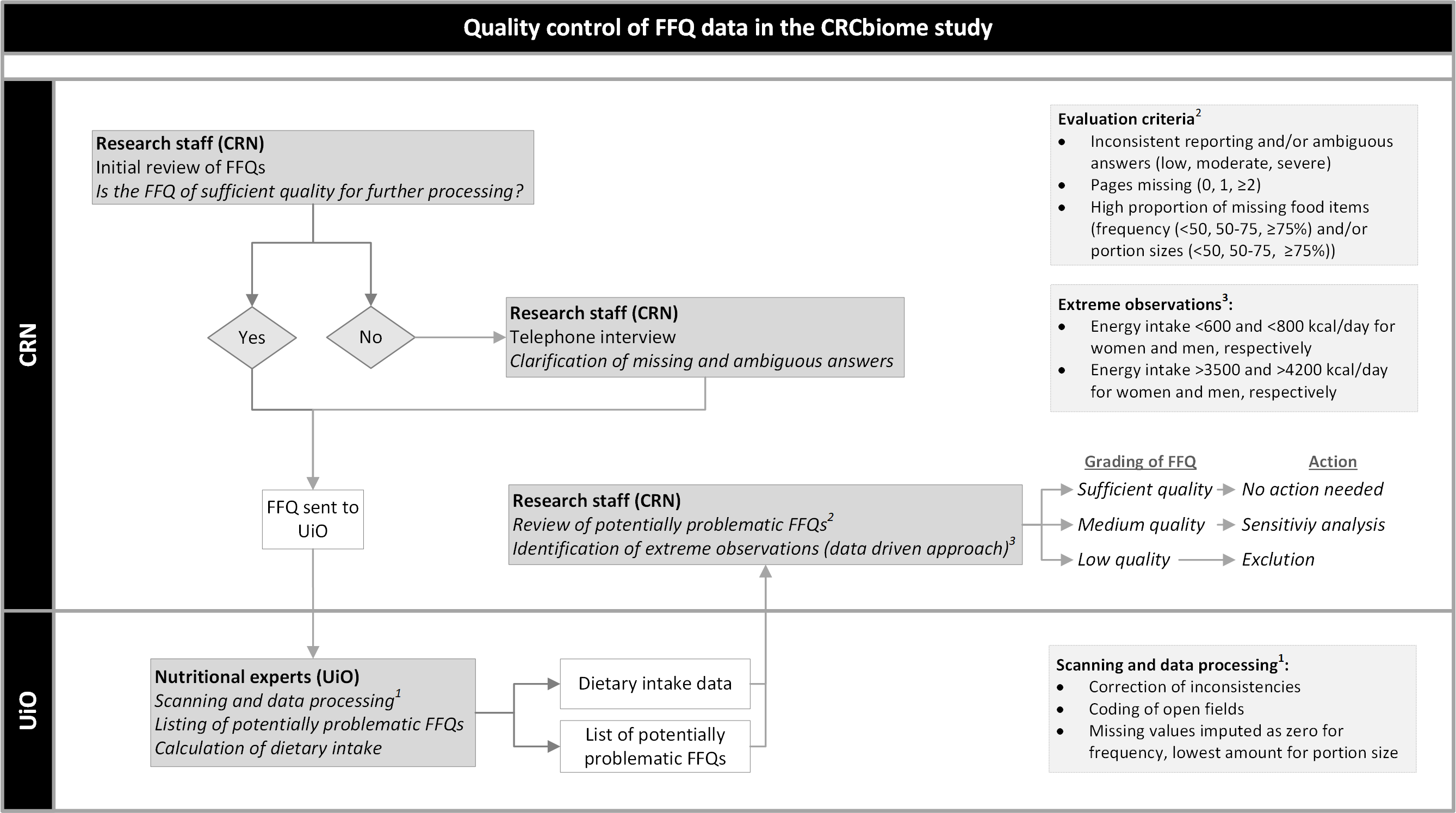

**Figure 1.** Upon receiving food frequency questionnaires (FFQs) from CRCbiome participants, completion is reviewed by researchers with expertise in nutritional epidemiology. Participants with FFQs of insufficient quality are contacted for clarification of inconsistencies and missing data. Reviewed questionnaires are then scanned using the Cardiff TeleForm program at the University of Oslo (UiO). Food and nutrient calculations are conducted using the software system KBS (“**K**ost**b**eregnings**s**ystem”/Dietary Calculation System) with the latest version of the food database, largely based on the Norwegian Food Composition Table (1). Missing answers are imputed as zero in line with common practice (2–5). Any FFQs regarded as potentially problematic during the data handling process are listed. Dietary intake data and the list of potentially problematic FFQs are then returned to the Cancer Registry of Norway (CRN). Potentially problematic FFQs are reviewed according to a set of predefined criteria, including inconsistency in reporting, number of missing pages and amount of missing food items. Based on these criteria, FFQs are graded as being of low, medium or sufficient quality. Whereas low quality FFQs will be excluded from all analysis where diet is the primary exposure, medium quality FFQs will be included unless sensitivity analysis indicates substantial attenuation of effect estimates. Lastly, in line with common practice in nutrition studies (6), observations with extreme energy intake levels in both the upper and lower range will be excluded.
