## Additional file 3 for "The CRCbiome study: a large prospective cohort study examining the role of lifestyle and the gut microbiome in colorectal cancer screening participants"

**Region:**
REC sør-øst D
**Contact person:**
Finn Skre Fjordholm
**Telephone:**
+47 22 84 58 21
**Date:**
18.12.2019
**Our reference:**
63148
**Your reference:**
Trine Ballestad B Rounge

**63148 The microbiome as a colorectal cancer screening biomarker**
The Cancer Registry of Norway
**Applicant**: Trine Ballestad B Rounge
**Applicants description of purpose:**

*Yearly, 1.4 mill people worldwide are diagnosed with colorectal cancer (CRC) and the incidence is increasing. Symptoms are unspecific - often emerging when the disease is no longer curable. Screening reduces CRC mortality, but current screening tests are either invasive, or hampered by poor sensitivity or specificity. There is a need for better tests. There is associations between gut microbiome and colorectal cancer. Lifestyle can affect the intestinal bacterial flora and cancer risk, but this interaction is little known. By mapping all the bacteria present in the gut one can develop tests that can be used to detect precursors and cancer early. Understanding the interactions between biomarkers for advanced neoplasia and lifestyle may guide measures for prevention of CRC and increase biomarker performance. Our main goal is to develop new tests for intestinal bacteria that can be used in future screening programs so that sampling is simplified and the result is more secure. We will also examine whether there is a connection between diet and lifestyle, gut microbiota and colorectal cancer development. Then we can improve advice on cancer prevention and increase the accuracy of the tests.*

**REC's assessment**
We refer to the application for prior approval of the above research project. The application was considered by the Regional Committee for Medical and Health Research Ethics (REC) south-east D) in the meeting 04.12.2019. The assessment is made based on the Health Research Act § 10. The project is a collection of two current sub-projects under REC 2011/1272 D «Pilot on a colorectal cancer screening program »and REC 2010/3087 A« S-98052a NORCCAP ».
All written inquiries about the case must be sent via the REC portal
You will find information about REC on our website RECportalen.no
It appears that the scope of the project has previously been approved by REC, and the application is shown to two decisions on change applications in REC 2011/1272 dated 17.3.2017 and 06.03.2018, and one decision in REC 2010/3087 dated 07.04.2016.

There is a connection between the gut microbiota and the risk of colorectal cancer, and the purpose of the project is to investigate this connection further. Participants must complete two

questionnaires before conducting the colonoscopy examination. This survey is part of the two previously approved projects, and data from this survey are used in this project.
Analyzes are made of a stool sample from REC 2011/1272. Furthermore, it must be handed in two stool samples in one year.
Summary information is obtained from the Cancer Registry and the Cause of Death Register. The Norwegian Prescription Database collects information on the use of antibiotics and medicines that affects the bowel. The committee has assessed the application and has no objections to the study as such. Committee has, however, several comments on the information letter and approves the project on terms that this is changed following these.

**Terms**
- It is stated in the information letter that the samples are stored «in a research biobank, together with the rest of the samples from Screening for bowel cancer - preliminary project ». The Committee assumes that this is about the biobank that is associated with REC 2011/1272. It is requested that it clarifies which biobank the sample is to be stored in and that the information sheet is updated so that the name of the biobank and the person responsible appear from the information letter.

- The information leaflet must contain more information about the project.
- In the introduction to the letter, the connection between the project and REC 2011/1272 should also and REC 2010/3087 are explained in more detail.

**Decision**
Approved with conditions REC has made a comprehensive research ethics assessment of all aspects of the project. The project was approved based on the Health Research Act § 10, provided that the above conditions are met.
All written inquiries about the case must be sent via the REC portal You will find information about REC on our website *REKportalen.no*
We also point out that according to the new Personal Data Act, there must also be one basis for processing following the Privacy Ordinance. It must be anchored in its institution. In addition to the conditions set out in this decision, the approval is given on the condition that the project is carried out as described in the application and protocol, and the provisions that follow from the Health Research Act with regulations.

The permit is valid until 01.01.2034. For documentation reasons, the information must nevertheless be preserved until 01.01.2039. The research file must be stored separately in a key and one information file. The information must then be deleted or anonymised, at the latest within one half a year from this date.
The research project's data must be stored properly, see the Personal Data Regulations Chapter 2, and the Norwegian Directorate of Health's guide for «Privacy and information security in research projects in the health and care sector ».
If significant changes are to be made to the project concerning the information provided is given in the application, the project manager must send a change notification to REC.
The project must send a final notification on a separate form, no later than six months after the end of the project.

The committee's decision was unanimous

Best regards

Find Wisløff
Professor em. Dr. med. , Leader

Find Skre Fjordholm, Adviser

Copy: Cancer Registry Administrative Leader/Director:

**Final message**
Applicants must send a final notification to REC south-east D on a separate form no later than six months after the approval period has expired, cf. hfl. § 12.
All written inquiries about the case must be sent via the REC portal You will find information about REC on our website RECportalen.no

**Application to make significant changes**
If you want to make significant changes in relation to purpose, method, time course, or organization, the application must be sent to REC that has given prior approval. The application must describe which changes are desired to be made and the reasons for these, cf. hfl. § 11.

**Right of appeal**
You can appeal against the committee's decision, cf. the Public Administration Act § 28 et seq. The appeal is sent to REC south-east D. The deadline for complaints is three weeks from the time you receive this letter. If the decision is maintained by REC south-east D, the complaint is forwarded to the National Research Ethics Committee for Medicine and Health Sciences (NEM) for final assessment.
