## Additional file 4A for "The CRCbiome study: a large prospective cohort study examining the role of lifestyle and the gut microbiome in colorectal cancer screening participants"

NORWEGIAN CANCER SOCIETY

Date: 30.10.2017

Dear Trine Rounge,

We are pleased to inform you that your grant application to the Norwegian Cancer Society's Open Call 2017 for the research project *The microbiome as a colorectal cancer screening biomarker* has been approved.

The total project grant for the period 01.07.2018 to 31.05.2022 is NOK 3500000. Please note that if your project receives only partial funding, the Norwegian Cancer Society may request a revised project proposal.

Your application has been carefully reviewed and considered by the peer review committee specified in your application. Grant applications are ranked on the basis of merit by one of five discipline-specific peer review committees, each of which is composed of 6 international researchers.

User representatives have taken part in the assessment process in each peer review committee. The aim of user representation is to assess whether user involvement is relevant and implemented in the project, and if the project has the potential of generating results with possible impact for patients.

An overview of RC 5: Epidemiological, health, and social science cancer research's evaluation of your proposal is provided below:

Relevance to cancer: Yes
Scientific quality: 6
Qualifications of the project manager and project group: 5
Impact: 4
Feasibility: 5

Overall score: 5.17 (considered fundable > 4.5)

The peer review committee´s comment to your proposal is provided below.

*This is a well-written application describing innovative and relevant research with potential for improving public health. However, we would like to see preliminary results and more detail about Substudy 1 before considering full funding. The stated aim of Substudy 1 is to "identify associations between the gut microbiota and advanced neoplasia in a Norwegian screening population" with "results are based on data from the target population" being promoted as a strength. The study, however, is performed in the subset (less than 20%) of the screening population who are FIT+. Is the exclusion of FIT- individuals made for logistic or scientific reasons, and what are the implications? If the aim is as stated, then we would like to see a motivation for using a selected sample and a discussion of how a selected sample (FIT+) will provide unbiased estimates for the target population (which includes FIT- individuals). The application lacks details of exactly how the new biomarkers will be used to improve screening.*

Please log into the Norwegian Cancer Society's web portal, go to *My tasks*, and indicate whether you wish to accept the awarded grant by **November 3, 2017**. Formal approval from the project administrator will subsequently be requested, upon which the Norwegian Cancer Society will send a formal contract to you and the project administrator.

Please note that you may not accept funding from two different sources for overlapping projects. In case of funding from both the Norwegian Cancer Society and another source, funding from the other source should be prioritized. In case of funding of two identical projects from two separate calls from the Norwegian Cancer Society within the same time period, you may only accept one grant.

General information about this year's call will soon be posted on our website. Please note that the right of appeal is limited to case handling errors and misuse of power, and must be received by the Norwegian Cancer Society within three weeks of receiving this letter. We kindly ask that questions regarding the evaluation process and/or the right to appeal be directed to.

Sincerely,

the Norwegian Cancer Society

Anne Lise Ryel

Secretary General

---

Post address: Postboks 4 Sentrum, 0101 Oslo

Visitation address: Kongens gate 6, Oslo

Internet address: www.kreftforeningen.no

CC: Giske Ursin
