## Additional file 4B for "The CRCbiome study: a large prospective cohort study examining the role of lifestyle and the gut microbiome in colorectal cancer screening participants"

NORWEGIAN **CANCER** SOCIETY

Date: 23.10.2018

Dear Paula Berstad,

We are pleased to inform you that your grant application to the Norwegian Cancer Society's Open Call 2018 for the research project *Gut microbiome and lifestyle for colorectal cancer risk classification (CRCbiome)* has been approved.

The total project grant for the period 01.01.2019 to 31.12.2022 is NOK 5600000. NOK 3600000 of the awarded grant comes from *Kreftforeningens paraplystiftelse for Kreftforskning*. Both the Norwegian Cancer Society and *Kreftforeningens paraplystiftelse for Kreftforskning* must be acknowledged as sources of finance in all publications related to the project.

Please note that if your project receives only partial funding, the Norwegian Cancer Society may request a revised project proposal.

Your application has been carefully reviewed and considered by the peer review committee specified in your application. Grant applications are ranked on the basis of merit by one of five discipline-specific peer review committees, each of which is composed of six international researchers.

User representatives have taken part in the assessment process in each peer review committee. The aim of user representation is to assess whether user involvement is relevant, and if so, to what degree it is implemented in the project.

An overview of committee 5: Epidemiological, health, and social science research's evaluation of your proposal is provided below:

Relevance to cancer: Ja
Scientific quality: 5
Qualifications of the project manager and project group: 6
Impact: 3
Feasibility: 5

Overall score: 5 (considered fundable > 4.5)

The peer review committee´s comment to your proposal is provided below.

*A well designed, large and realistic project with potentially important results. However, given the diifficulty in measuring lifestyle, especially food habits across long time periods, we would have liked to see a better description of the validity of the lifestyle and food questionnaires, in line with what was written for the other proposed methods. The application seems to reflect work carried out by a large croup of investigators where it is not entirely clear what role the applicant will have. There are some plans for user involvement, but it is vague and inadequately described. This should be improved.*

The awarded grant must be accepted by your project administrator. You will receive a copy of the formal contract between Kreftregisteret and the Norwegian Cancer Society.

Please note that you may not receive funding from two different sources for overlapping projects. In case of funding from both the Norwegian Cancer Society and another source, funding from the other source should be prioritized. In case of funding of two projects from two separate calls from the Norwegian Cancer Society within the same time period, you may only receive one grant.

General information about this year's call will soon be posted on our website. Please note that the right of appeal is limited to case handling errors and misuse of power, and must be received by the Norwegian Cancer Society within three weeks of receiving this letter. We kindly ask that questions regarding the evaluation process and/or the right to appeal be directed to.

As a recipient of research funding, we welcome you to this year's award ceremony. The ceremony will take place on Tuesday October 30 from 12.30-14.30 at the Norwegian Cancer Society's Science Centre, located in Kongens gate 6, Oslo. We would very much like you to be present at this event!

Please register to attend here by Thursday October 25 at 16.00.

Sincerely,
The Norwegian Cancer Society

Anne Lise Ryel
Secretary General
-----------------------------------------------------
Post address: Postboks 4 Sentrum, 0101 Oslo
Visitation address: Kongens gate 6, 0153 Oslo

www.kreftforeningen.no

CC: Giske Ursin
