## Additional file 4C for "The CRCbiome study: a large prospective cohort study examining the role of lifestyle and the gut microbiome in colorectal cancer screening participants"

Hi all

Congratulations on the allocation of research funds from HSØ 2020!

The board of Helse Sør-Øst RHF has allocated NOK 134 million to new research projects and research networks for 2020.

https://www.helse-sorost.no/nyheter/tildeling-av-regionale-forskningsmidler-for-2020

Your / your new project (s) must have a project number and be linked to the correct costCODE, ie where salaries and purchases of goods and services are to be booked. This is important so that you can follow the use of the funds, as well as be able to report during the project phase.

To start the process with the project number, you must answer the email sent from Helse Sør-Øst to confirm that you wish to receive an award, and in addition confirm that there is no double financing of your project. You can also inform about when the start-up of the new project is expected.

When this has been done, you must contact financial resources in your own clinic to clarify which cost code is to be used for your project and they will also apply for a project number for you via Forskningsstøtte's EFP (Externally Funded Projects) register.

NB! These are funds allocated to OUS-HF, and cannot be transferred to another institution - allocations with the project manager at the Cancer Registry or another institution must remember to state the cost coe for the cooperating unit at OUS for employment and placement of the funds.

Assignment of project number will be sent to you after final confirmation from Health South-East in early January. Therefore nice if you confirm to HSØ on your inquiry as soon as possible if you want to receive the funds. Then the process starts faster here too internally.

Wishing you a very MERRY CHRISTMAS and a HAPPY NEW YEAR!

Sincerely, Research Support;

v / Trine-Lise Grimsrud
